# A vulnerability index for COVID-19: spatial analysis to inform equitable response in Kenya

**DOI:** 10.1101/2020.05.27.20113803

**Authors:** Peter M Macharia, Noel K Joseph, Emelda A Okiro

**Author notes:** joint first authors (Peter M Macharia, Noel K Joseph).

## Abstract

**Background:** Response to the COVID-19 pandemic calls for precision public health reflecting our improved understanding of who is the most vulnerable and their geographical location. We created three vulnerability indices to identify areas and people who require greater support while elucidating health inequities to inform emergency response in Kenya.

**Methods:** Geospatial indicators were assembled to create three vulnerability indices; social (SVI), epidemiological (EVI) and a composite of the two (SEVI) resolved at 295 sub-counties in Kenya. SVI included nineteen indicators that affect the spread of disease; socio-economic inequities, access to services and population dynamics while EVI comprised five indicators describing comorbidities associated with COVID-19 severe disease progression. The indicators were scaled to a common measurement scale, spatially overlaid via arithmetic mean and equally weighted. The indices were classified into seven classes, 1-2 denoted low-vulnerability and 6-7 high-vulnerability. The population within vulnerabilities classes was quantified.

**Results:** The spatial variation of each index was heterogeneous across Kenya. Forty-nine north-western and partly eastern sub-counties (6.9 m people) were highly vulnerable while 58 sub-counties (9.7 m people) in western and central Kenya were the least vulnerable for SVI. For EVI, 48 sub-counties (7.2 m people) in central and the adjacent areas and 81 sub-counties (13.2 m people) in northern Kenya were the most and least vulnerable respectively. Overall (SEVI), 46 sub-counties (7.0 m people) around central and south-eastern were more vulnerable while 81 sub-counties (14.4 m people) that were least vulnerable.

**Conclusion:** The vulnerability indices created are tools relevant to the county, national government and stakeholders for prioritization and improved planning especially in highly vulnerable sub-counties where cases have not been confirmed. The heterogeneous nature of the vulnerability highlights the need to address social determinants of health disparities, strengthen the health system and establish programmes to cushion against the negative effects of the pandemic.

**Summary:** *Key questions:*

What is already known?
- Disasters and adverse health events such as epidemics and pandemics disproportionately affect population with significantly higher impacts on the most vulnerable and less resilient communities.
- Significant health, socio-economic, demographic and epidemiological disparities exist within Kenya when considering individual determinants, however, little is known about the spatial variation and inequities of their concurrence. What are the new findings?
- Sub-counties in the north-western and partly eastern Kenya are most vulnerable when considering social vulnerability index while central and south-east regions are most vulnerable based on the epidemiological vulnerability index affecting approximately 6.9 million and 7.2 million people respectively.
- The combined index of social and epidemiological vulnerabilities shows that on average, 15% (7.0 million) of Kenyans reside in the most vulnerable sub-counties mainly located in the central and south-eastern parts of Kenya. What do the new findings imply?
- Targeted interventions that cushion against negative effects to the most vulnerable sub-counties are essential to respond to the current COVID-19 pandemic.
- Implementation of strategies that address the socioeconomic determinants of health disparities and strengthening health systems is crucial to effectively prevent, detect and respond to future adverse health events or disasters in the country.
- Need for better quality data to define a robust vulnerability index at high spatial resolution that can be adapted and used in response to future disasters and adverse health events in the long run.

## Introduction

There has been growing recognition of the threat that epidemics, disasters and public health emergencies pose to global health security and the livelihoods of people, beyond their impact on human health [1]. Under the umbrella of the global health security agenda, countries have come together to advance a world safe and secure from infectious disease threats. Hence, countries are expected to be prepared to weather disease outbreaks and natural disasters with Africa having a history of dealing with emerging diseases [2,3] most recently the Ebola epidemic in West Africa and DRC [4].

Epidemics, disproportionally impact vulnerable populations as can be witnessed with the coronavirus disease 2019 (COVID-19) in the United States and the United Kingdom where Black, Asian and minority ethnic communities are being disproportionately affected [5-7] as are poorer communities [8,9]; during the Ebola outbreak the disease was linked to rural and remote areas [10]. The reasons behind this disparity are complex and varied however, one of the biggest underlying factors driving this disproportionate impact is socioeconomic inequities [8,11]. Several factors, including poverty, lack of access to healthcare and transportation, and certain aspects of housing may weaken a community’s ability to prevent significant human and financial loss when a disaster such as the current COVID-19 pandemic strikes. A framework that provides a detailed understanding and location of population groups that are either vulnerable to increased risk of infection or inferior health outcomes and that are also marginalized from health services can inform where additional resources are most needed. These factors are collectively identified as vulnerability and aim to identify people that are disproportionally exposed to the risk of infection and/or disease severity.

COVID-19 rapidly spread worldwide and while low testing numbers do not allow for a reliable appraisal of the extent of the epidemic in Africa, 115,616 cases and over 3,479 deaths had been reported in Africa as of May 26^th^, 2020 [12]. Health officials expect that it is only a matter of time before infections begin to rise in Africa with growing concerns about risks in a continent that faces unique challenges in dealing with the current outbreak [2]. WHO has estimated that between 83,000 and 190,000 people could die of COVID-19 and 29 million to 44 million could get infected during the first year of the pandemic if containment measures fail in Africa [13]. However, although cases are increasing, most countries in Africa are not documenting the same exponential growth rate in confirmed cases as seen in Europe and the United States [11,14].

The first case of COVID-19 in Kenya was confirmed on March 13^th^, 2020. Since then 1,348 cases have been reported with 52 deaths confirmed by May 26^th^ 2020 [15]. The Kenyan government, through the Ministry of Health, has responded quickly by taking proactive public health measures to combat the spread of the disease. These measures have been through intensive tracking of contacts, isolation of confirmed cases, halting non-essential services, and cutting off routes of transmission through suspension of international flights, dusk-to-dawn curfew, partial lockdowns and most recently closing borders with high intensity neighbouring countries of Tanzania and Somalia [15]. However, certain challenges face the implementation of some of these measures. There has been widespread growth of informal settlement in Kenya which typically are characterized by high-density neighbourhoods and poor infrastructure common across the continent where self-isolation and social distancing are proving extremely challenging. A substantial number of people work in the informal sector [16] where working from home is not realistic and the risks of a sudden loss of income is a constant threat with many more suffering from insecure food supplies [17].

Often people living in these settings often have malnutrition, infectious diseases such as HIV/AIDS and some may suffer from non-communicable diseases putting these individuals at greater risk of more severe clinical infection and mortality due to COVID-19 [18]. Other challenges include high poverty rates, weaker health systems and minimal access to healthcare with most countries having inadequate financial leeway to provide an effective response without external assistance. While the scarcity of resources is a matter of concern, it is equally important to look at whether the resources available support the most affected communities. This time presents an opportunity to reduce overall inequities by supporting the most vulnerable groups. Some level of prioritization and targeting of resources will prove useful in this context to identify those people and places that face the highest risk.

Using a wide range of spatially referenced indicators to enumerate a varied range of social constraints and assess risk, we developed three COVID-19 specific vulnerability indices to enumerate both social vulnerability (affects the risk of infection and spread) and epidemiological vulnerability (affects the risk of progression to severe disease) defined at the sub-county level in Kenya. This will facilitate identification of vulnerable groups and elucidate health inequities to help public health response by identifying and mapping communities that will most likely need greater support during the current outbreak.

## Data and methods

Geographic analysis unit: In 2013, Kenya adopted a decentralized system of governance where the units of administration and health planning were revised to 47 counties with broad policy directions maintained at the national level [19,20]. The counties are further sub divided to sub-counties [21] and was adopted as the unit of analysis. A geospatial layer of all sub-counties was created by digitizing sub-county maps available for each county from the county integrated development plans (CIDPs) [21]. Additional file 1 shows all the 295 sub-counties of Kenya derived from the CIDPs.

Three indices were defined, social vulnerability index (SVI), epidemiological vulnerability index (EVI) and a composite of the two, the social-epidemiological vulnerability index (SEVI). SVI broadly refers to the resilience of communities in face of external stresses such as disasters and disease outbreaks on human health [22]. The indicators used to define SVI entail socio economic inequalities [17,23,24], population dynamics [23] and access to services [23-26] which are likely to affect the risk of infection and spread of the disease at different rates in different sub national areas.

The EVI encapsulates diseases and comorbidities that affect the likelihood of disease progression hence affecting the severity of COVID-19 disease [18,27-30]. They include underlying medical conditions such as liver disease, chronic kidney disease, obesity, hypertension, diabetes, smoking, cardiovascular disease, chronic respiratory disease, cancer and people who are immunocompromised including HIV/AIDS [18,27-30]. The SVI and EVI were combined to generate SEVI in order to explore the overall resilience and risk of developing severe COVID-19 disease in Kenya.

### Data assembly

To create the indices at subnational level in Kenya, 24 data layers were assembled from various sources (Table 1) based on data availability, Kenya’s epidemiological context and preliminary findings suggesting an association with COVID-19 disease [17,18,23-30]. Nineteen datasets were used to define the SVI while five were used to construct the EVI. The datasets were available in different formats and at different spatial resolution necessitating pre-processing and/or modelling before being input to the framework for constructing a vulnerability index.

**Table 1:**
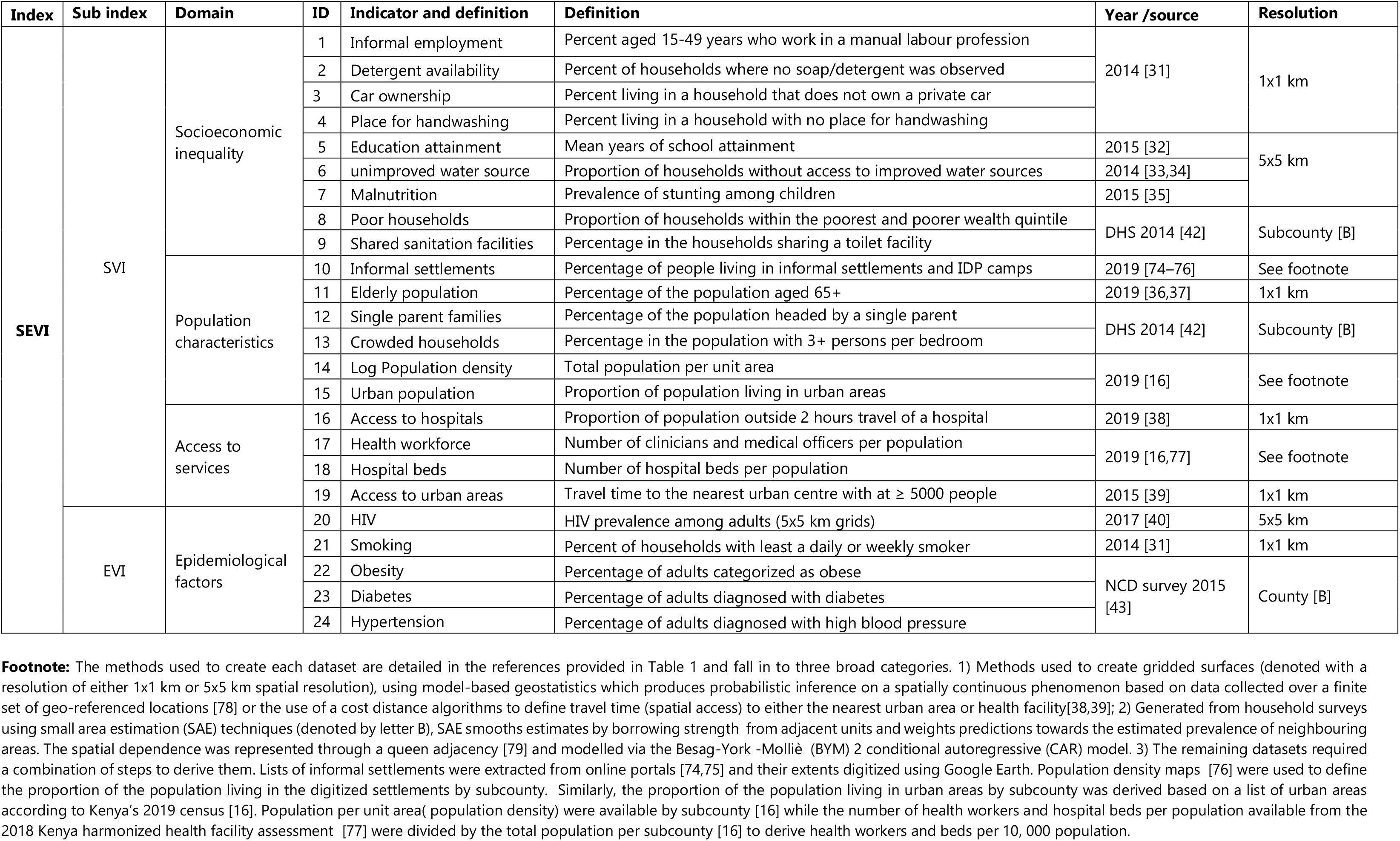
Data layers used to define COVID-19 vulnerability index in Kenya including their definition, sources, spatial resolution, year of they refer to and pre-processing done. The layers are classified into 4 thematic areas

The 24 layers of data fall into three broad groups. The first group comprised of twelve data layers available as gridded surfaces at 1 × 1 km or 5 × 5 km spatial resolutions [31-40] [Table 1]. The mean value per indicator and subcounty was extracted using the *zonal statistics* function of the Spatial Analyst tool of ArcMap 10.5 (ESRI Inc., Redlands, CA, USA). The second group comprised of seven variables whose coverage was estimated using small area estimation (SAE) models [41] based on the 2014 demographic and health survey (DHS) [42] and the 2015 stepwise survey for non-communicable diseases (NCD) [43] conducted in Kenya [Table 1 denoted as B]. DHS 2014 and NCD survey were designed to provide precise estimates at county and provincial level respectively. Therefore, SAE models were used to smooth estimates at subcounty (DHS 2014) and county level (NCD survey) using R-INLA package [44] in R software (version 3·4·1). The remaining five data layers were derived through a combination of geospatial techniques (Table 1 footnote).

### Constructing SVI

COVID-19 disease vulnerability indices have been created using different approaches ranging from technical [45-47] to simpler approaches [48-50]. In the current analysis, the indices were created following approaches used to define universal health coverage (UHC) and equity in maternal and child health [51-55], infectious disease vulnerability index [56] and some of the recent COVID-19 vulnerability indices [48-50].

To create SVI for Kenyan sub-counties, three sub domain indices were first defined based on major thematic areas related to COVID-19 vulnerability (Table 1 and Figure 1). They included socioeconomic inequality, population characteristics and access to services. The subdomains facilitate a finer and detailed viewpoint that would have been masked by a single averaged index. A similar approach was used when creating an infectious disease vulnerability index for African countries [56] and a composite coverage index for measuring UHC [53].

Nine indicators (Table 1 and Figure 1) were used to define socioeconomic inequality sub domain. People of low socioeconomic status including those under casual employment and physical labour are unlikely to have resources to actualize measures put in place by the government such as social distancing viz-a-viz earning a daily livelihood [7,17,24]. Further, households need to have access to a basic handwashing facility, soap and detergents as a first line of protection against infection [17]. The population characteristics subdomain included six indicators related to population dynamics (Table 1 and Figure 1). As population density increases, the rate of transmission of infectious diseases increases [57], informal settlements, refugee and internally displaced people (IDP) camps are vulnerable mainly due to shared community facilities such as water points and toilets while rural areas maybe at a lower risk [58]. The risk of COVID-19 increases with age especially after sixty years [17,18,59].

Finally, four indicators were used to define access to services (Table 1, Figure 1). About 8% of the Kenyan population lives outside two-hour travel of the nearest facility capable of offering hospital and emergency care [38] that might be needed to deal with severe COVID-19 cases with significant gaps in hospital capacity to accommodate a surge due to COVID-19 [26]. Further, commodities and services that might be needed by people during the COVID-19 era are likely to be concentrated in urban areas with marginalized areas associated with poorer health and education outcomes [39].

The geospatial data layers in each subdomain had different scales with different minima and maxima values. Therefore, to make the values comparable, they were first rescaled to a common scale ranging between 0 (least vulnerable) and 100 (most vulnerable) (Equation 1). The scaled indicators were used to create a unique index for each of the three subdomains through an arithmetic mean and were equally weighted. The arithmetic mean of the three subdomains indices was used to create the SVI.

**Equation 1:** Rescaling of coverage and or prevalence values for each determinant to a common scale ranging from 0 (least vulnerable) to 100 (most vulnerable)

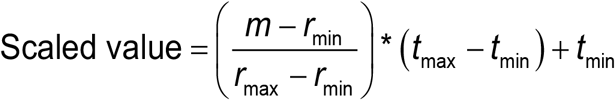

M - the value to be scaled; r min- the minimum value in the original range; r max- the minimum value in the original range; t min - the minimum value in the new scale (0) and t max - the maximum value in the new scale (100)

### Constructing EVI and SEVI

Five indicators on underlying health conditions and comorbidities, risk factors for severe disease [18,27-30] were available including the prevalence of HIV, smoking, obesity, diabetes and hypertension (Table 1, Figure 1). Similarly, these layers were first rescaled (Equation 1) and spatially overlaid using arithmetic mean and equally weighted to generate EVI. Finally, the generated SVI and EVI were averaged to generate the composite final index, SEVI.

Each index was grouped into seven classes ranging from the least vulnerable to the most vulnerable. The classes were grouped using the natural Jenks classification method which identifies “natural” groups within the data by reducing the variance within classes and maximizing the variance between classes [60]. A higher group (the sixth and seventh class) indicated high vulnerability while a lower group (the first and the second class) represented low vulnerability. The groups are unique within each index since the distribution of data among the indices are different. The proportion of the population within each band of vulnerability was then computed based on the 2019 Kenya’s census data.

Figure 1 summarizes the input data and methods used to define the indices in Kenya across the 295 sub-counties. The analysis and visualizations were done using StataCorp. 2014 [Stata Statistical Software: Release 14. College Station, TX: StataCorp LP], R software (version 3·4·1) and ArcMap 10·5 (ESRI Inc., Redlands, CA, USA).

**Figure 1:**
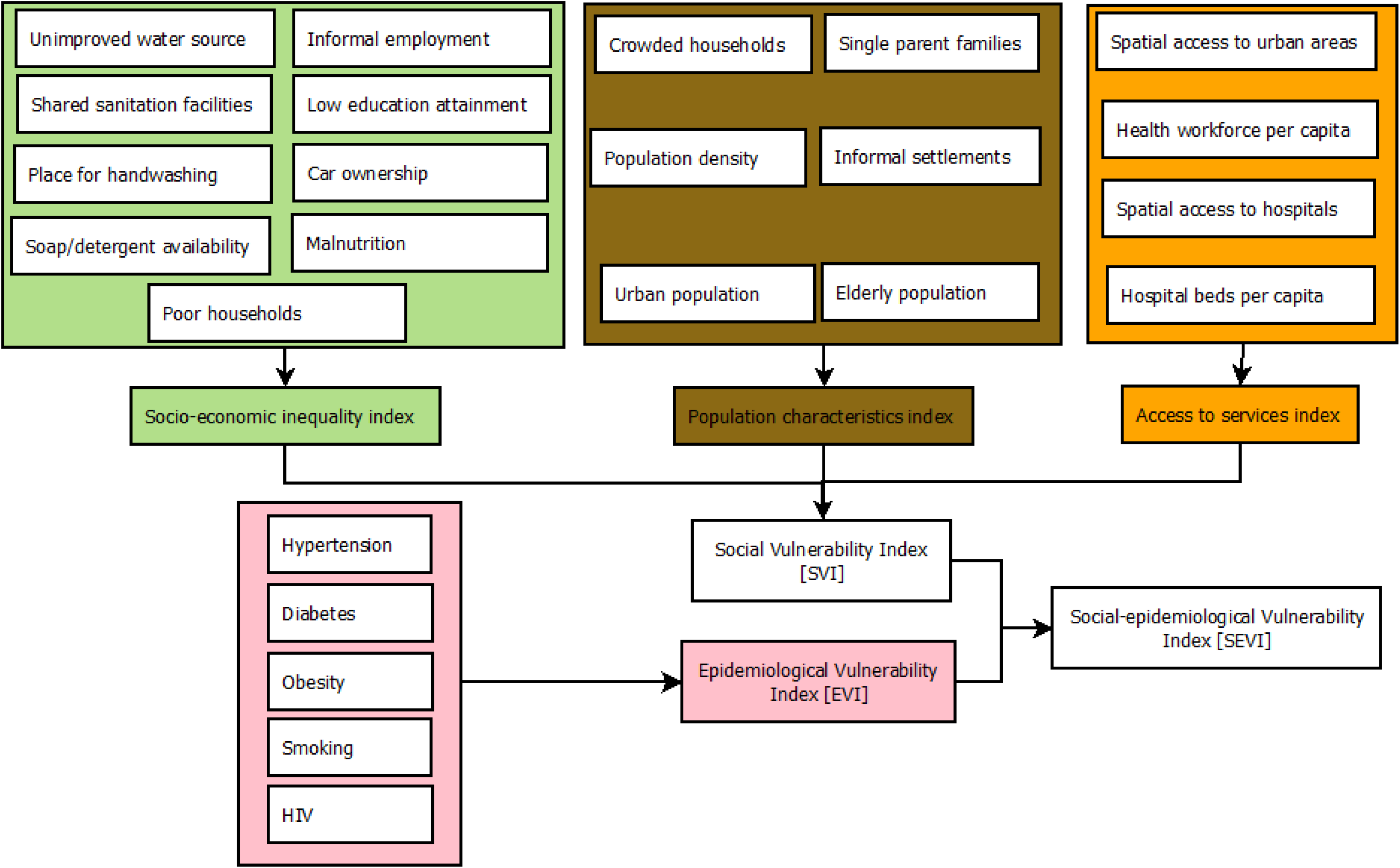
Schematic representation of data layers and approaches used to define social vulnerability index (SVI), epidemiological vulnerability index (EVI) and the combination of the two, social-epidemiological vulnerability index at subnational level in Kenya.

### Patient and public involvement

Our study does not involve the participation of patients or any members of the public. All data used in this study are aggregated and publicly available as listed in Table 1.

## Results

The spatial variation of the social vulnerability index (SVI) was heterogeneous across the 295 sub-counties (Fig 2A). The least vulnerable sub-counties are mainly located in the western and central parts of Kenya while sub-counties in north-eastern and southern parts are moderately vulnerable. North-western and parts of eastern Kenya have the most vulnerable sub-counties (Figure 2A). Approximately fifteen per cent (6.9 million) of Kenya’s total population resides in the 49 sub-counties that that were classified as highly vulnerable. Almost two-thirds (65%) of Kenya’s population (30.9 million) live in 188 sub-counties that are moderately vulnerable while only circa 21% live in the 58 least vulnerable sub-counties.

The most vulnerable sub-counties in north-western and partly eastern Kenya with reference to SVI are characterized by poor geographic access to health care services, marginalized in terms of access to the nearest urban areas and economically disadvantaged (mainly poor households, poor access to improved water and sanitation and low education attainment) (Additional file 2, and Additional file 3). However, despite being the most vulnerable, the region has a low population density and fewer families with single parents relative to other parts of Kenya (Additional file 2).

The least vulnerable sub-counties based on SVI mainly in central and western Kenya have a lower proportion of the poor households, improved access to water and handwashing soaps/detergents, higher education attainment while most settlements are near urban areas and within 2 hours of the nearest hospital (Additional file 3). Despite having on average low vulnerability, both central and western Kenya sub-counties have a high population density and higher proportion of the elderly population. In addition, sub-counties in central Kenya and adjacent areas have a slightly higher number of urban population and families with a single parent (Additional file 2).

**Figure 2:**
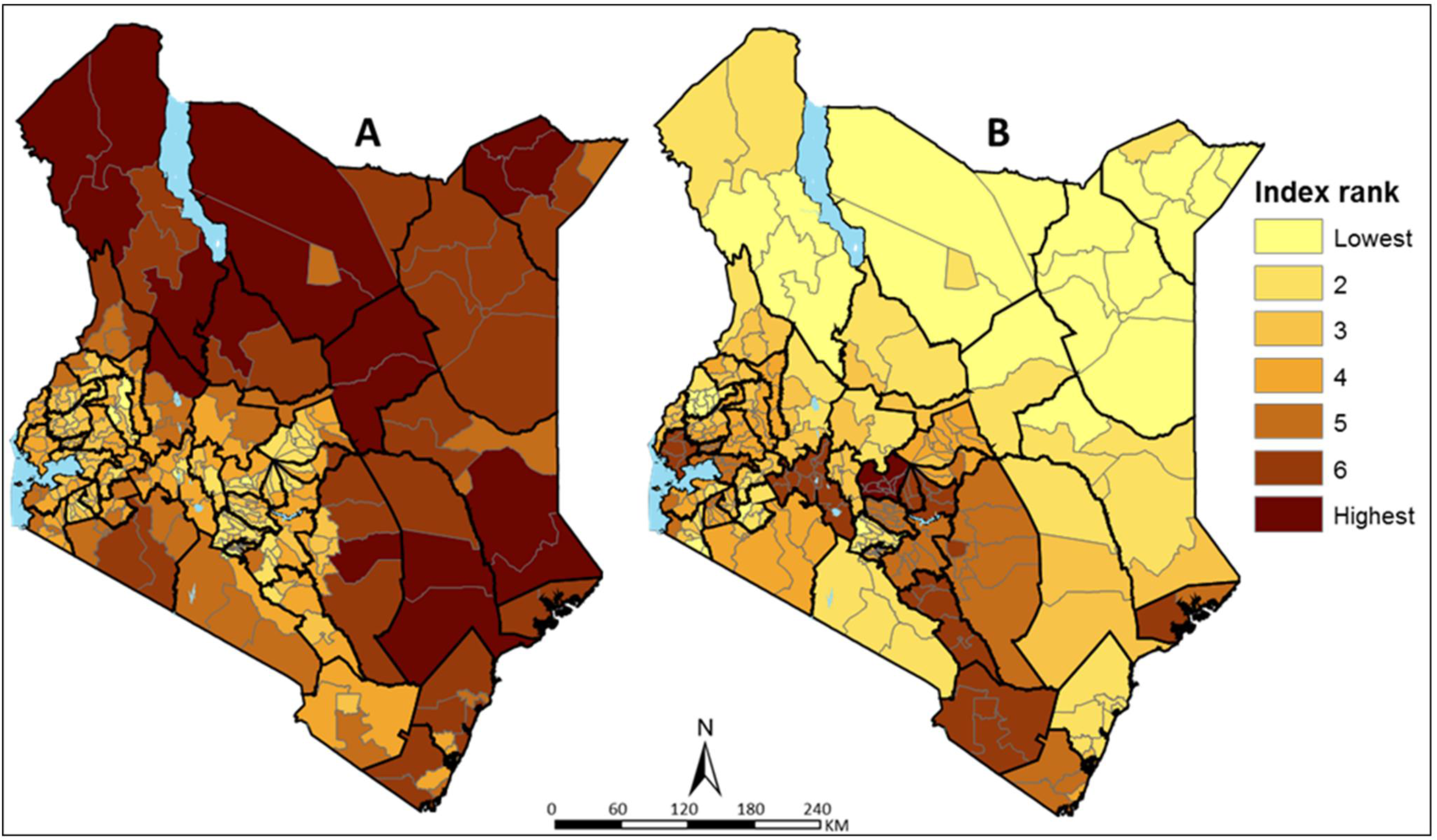
Social vulnerability index (SVI) (A) and epidemiological vulnerability index (EVI) (B) across 295 sub-counties in Kenya grouped into seven ranks. Rank 1 and 2 are the least vulnerable sub-counties while rank 6 and 7 are the most vulnerable.

Conversely, based on the EVI, 48 sub-counties in central, south-east and partly western Kenya where approximately 7.2 million people reside were the most vulnerable (Figure 2B). Approximately thirteen million (13.2) people residing in 81 sub-counties mainly located in the northern and eastern parts were classified as the least vulnerable. Majority of the sub-counties in the south of the equator have higher epidemiological vulnerability mainly driven by a high prevalence of hypertension and smoking. High prevalence of obesity and diabetes is only evident in fewer sub-counties around central and south-east Kenya while HIV is more prevalent in the western Kenya (Additional file 2, and Additional file 3). There are exceptions in north-eastern, where Wajir county has a higher prevalence for both diabetes and hypertension while Turkana county has a higher prevalence of smoking (Additional file 2).

The spatial variation of the overlay between social vulnerability and epidemiological vulnerability (SEVI) is shown in Figure 3. The resulting patterns are smoother than EVI and SVI since its average of the two which were had fewer areas of concurrence of the high or low vulnerable sub-counties. Consequently, majority of Kenya’s population (55%) reside in moderately vulnerable sub counties mainly in north-western and eastern parts of Kenya.

Forty-six sub-counties in the central and adjacent areas, south-east and partly western Kenya were the most vulnerable affecting 15% (7.0 million) of Kenya’s population. More importantly, a few sub-counties that are highly vulnerable due SEVI, have a dual burden of SVI and EVI. Notable cooccurrence of EVI and SVI include parts of Kitui (central-east), Elgeyo Marakwet (partly western) and Narok (south-west). These areas have a high prevalence of smoking, hypertension and stunting, higher proportion of elderly population, low access to improved water and sanitation and smaller proportion of people within 2-hours of the nearest hospital.

Approximately 30% of the population reside in sub-counties classified as the least vulnerable based on SEVI. These sub-counties are not localized in one geographic but scattered across Kenya mainly in western (e.g. Bungoma county) and a few in north-eastern (e.g. Wajir county) and central (e.g. Kiambu county) Kenya. The localized areas of low vulnerability have different variable factors contributing to the least vulnerable score. For example, in Kiambu, central Kenya, all 24 indicators have low scores (meaning least vulnerable) except five, namely, high population density, high urban population, elderly population, shared toilets and high prevalence of smoking whose effect is masked in the combined index. On the other hand, Wajir county in north-eastern, only crowded and poor households, poor spatial access to hospitals, water and sanitation had higher scores (meaning more vulnerable) whose effect was neutralized by low scores from the rest of the indicators.

**Figure 3:**
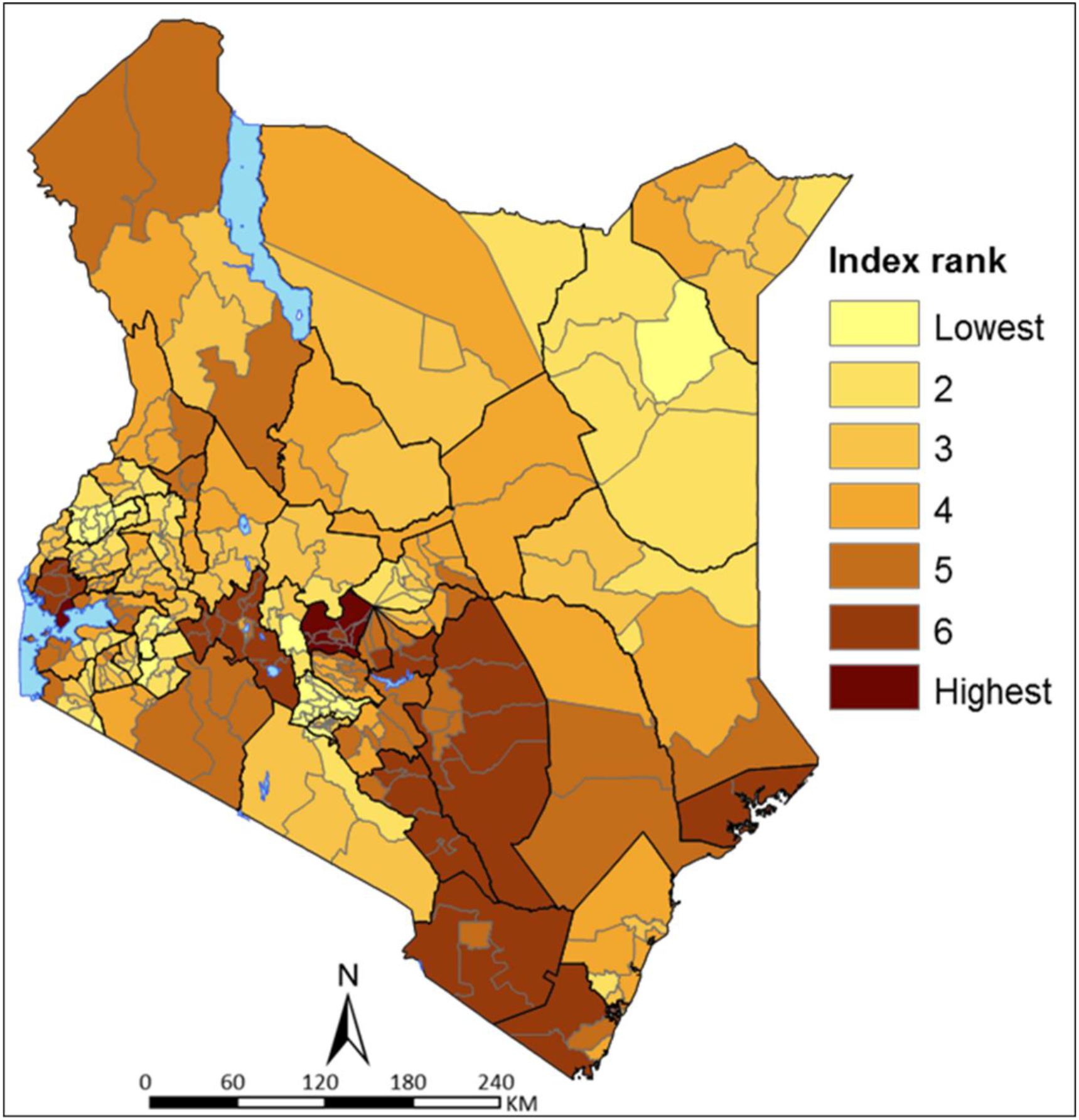
Social- epidemiological vulnerability index (SEVI) across 295 sub-counties in Kenya grouped into seven ranks. Rank 1 and 2 are the least vulnerable sub-counties while rank 6 and 7 are the most vulnerable.

## Discussion

Emerging and re-emerging diseases with pandemic potential continue to challenge countries and health systems, causing enormous human and economic losses [1,3]. Health security calls for the need for all countries to invest in improving their global health preparedness in the phase of emerging epidemics including stronger health infrastructure as the best defence against disease outbreaks and other health threats [2,61]. Not surprisingly most countries are struggling to mitigate the impact of the current COVID-19 pandemic with varied levels of success with increasing fears or re-emergence in those places that have managed to contain the pandemic [62]. It remains uncertain how Africa is going to come through this pandemic. Compared with the rest of the world, the virus has arrived later, providing an opportunity to learn from other contexts that could help guide Africa’s fight. Social distancing and basic hygiene measures are proving to be the most effective tools to slow down the rate of transmission. Yet, in many contexts throughout Africa, social distancing and frequent handwashing are privileges that not everyone has access to [7,63].

In Kenya, the expectation is that nearly all communities will be affected by COVID-19 to yet undetermined degrees; however, the impact of the pandemic will not be the same in each locality. Meanwhile, Kenya does not have a preexisting granular vulnerability index such as the CDC’s US social vulnerability index [22]. Such an index would have been used to assess vulnerability or identify vulnerable populations that can be used to inform decisions on the disbursement of social support measures or identify those areas that require improved health services. In the current analysis three indices have been developed to identify which communities may need the most support as COVID-19 spreads in the country. Mapped to the sub-county, the vulnerability indices provide information that is useful for emergency response planning and mitigation at a relatively granular level and can help support response planning for the current epidemic.

Once introduced, outbreaks spread faster in vulnerable communities than in less vulnerable areas. Some communities in Kenya were identified to be more vulnerable than others and would exhibit compromised ability to manage the spread and limit the economic and social impact of the outbreak. The social vulnerability index identified sub-counties in the northern and south east parts of Kenya as the most vulnerable while majority of the sub-counties in central and western Kenya were observed to be less vulnerable (Figure 2A). The 49 most vulnerable sub-counties account for 6.9 million (15%) of Kenya’s total population require greater focus and prioritization in terms of response.

There is a divergence between sub-counties that had high social vulnerability indices and those defined as epidemiologically most vulnerable (Figure 2). Most of the sub-counties in western, central and parts of south east were observed to have high EVI hence when applied to COVID-19, these areas are likely to have sub populations at higher risk of developing severe disease and increased mortality rates [18,27-30]. While the Kenyan population has hypertension, obesity and diabetes, the prevalence is generally not high compared to other settings where severe disease and increased mortality have been observed. Conversely these regions appear to be less vulnerable with respect to social vulnerability (Figure 2A).

Though sparsely populated, northern and south eastern parts of Kenya were less epidemiologically vulnerable and are therefore generally less vulnerable to severe diseases but more susceptible to infections and spread when considering their socioeconomic context. They have poor access to hospitals and urban areas, high number of poor households, constrained access to water and sanitation and low education attainment. These metrics allow for identification of geographic areas that are most likely to harbour large numbers of undocumented COVID-19 cases due to lack of access to care (Additional file 2, Additional file 3), which in turn can inform the geographic targeting of testing and surveillance efforts and for the deployment of temporary hospitals based on projected need.

The government of Kenya has put in place several measures to curb the spread of COVID-19. Some of these measures lead to reduced social interaction hence reduced production and demand across all the sectors which are costly to the economy and will have negative impacts on the livelihoods of people. The national government, the county governments and other stakeholders are implementing programmes to cushion against adverse socio-economic impacts. These include reduction of taxes, provision of masks and hand sanitizers, distribution of food, water and other commodities. Further the government will inject Kenya shillings 53.7 billion into the economy to stimulate growth and cushion families and companies during COVID-19 pandemic in eight thematic areas including infrastructure, education, small and medium enterprises, health, agriculture, tourism, environment and manufacturing [64]. This analysis identifies areas that should be prioritized for different interventions when programmes to ease vulnerability and mitigate the effects of COVID-19 are being rolled out across the country. This is important especially in areas that are highly vulnerable and are yet to experience an escalation of cases. These indices have the potential to pave the way for data driven informed planning to tackle vulnerability.

The epidemic is shaped by many factors, testing capacity and social distancing, as well as population density, age structure, wealth and other social behavioral factors. In Kenya the spread of the virus is uneven with most of the cases identified in the capital city, larger towns and a few border towns. By overlaying the number of confirmed cases of COVID-19 onto the vulnerability indices, we can begin to explore the spread of the virus in communities with different levels of vulnerability. We have preliminary explored how vulnerability relates to the numbers of confirmed cases of COVID-19 using case data at the county level made available by of May 14th.Identified hotspots (Mvita subcounty, Dagoretti north and Kamukunji sub-counties) are in highly vulnerable areas when considering their population characteristics which include being largely urban, high population density with a high proportion of people living within informal settlements. Further they have most people within informal employment and shared sanitation facilities. This is somewhat confounded by the fact that we are unable to separate out imported cases which comprise a substantial percent of the total case count.

Countries are starting to ease lockdown measures to limit the negative impact on the economy [62,65]. These decisions need to be informed by the trends in new cases, the potential risk of resurgence and the strength of public health systems including the capacity to detect new cases. The vulnerability index delivers a measure through which to better appreciate factors that enable communities to remain resilient, inform on their ability to carry out personal protective measures, practice both hand hygiene and hygiene in the household and the possibility of social distancing in different context. Importantly these indices have identified indicators that shed light on factors that would drive the continued spread of disease and inform prediction on the burden of severe disease and mortality due to COVID-19 in Kenya. Importantly, the indices developed are versatile and can be repurposed for use in varied emergency response situations.

The indices showed widespread inequities across sub-counties of Kenya. While the interim measures will help ease the pressure and reduce vulnerability, there is a need to reduce inequities in the longer term, beyond the current COVID-19 pandemic in preparation for future epidemics which are inevitable [66]. Africa must invest heavily in relevant data systems and preparedness by increased government investments. Programmes to ensure access to improved water and sanitation, targeted social programmes such as raising awareness for proper hygiene, improvement of housing facilities in the informal settlements and IDP camps are needed. Strengthening health systems remains at the core of reducing health inequities [66]. The system should also be redesigned to deal with surge capacity by absorbing the increase in the demand of healthcare services due to epidemics and pandemics [26].

The prevalence and recurrence of epidemics and disasters in Africa should be the impetus for greater investment in preparedness. Disasters such as COVID-19 pandemic [67,68], Ebola epidemic [4], flooding in East and Southern Africa [69], ongoing floods and recurrent malaria epidemics in Kenya [70] have become common. Investments on measures such as early warning systems should be put into place to detect, respond and effectively contain these threats. Strategic actions that were recommended against influenza could potentially inform better preparation in case of a viral disease; capability to develop pandemic strain vaccines, stockpiles of broad-spectrum antiviral drugs, surge capacity for rapid vaccine production and developing models that could inform effective means of delivering therapies during an outbreak [71].

The indicators used in the current analysis to derive vulnerability indices were based on the priori understandings of who is a risk and vulnerable from information available so far, however, COVID-19 is evolving and more insights will become increasing available and the indices can be adapted. Further the classification of indicators into sub domains should be adapted to context and the data availability. For example, Wilkinson *et al*.,(2020), proposes categorizing the indicators into epidemiological vulnerability (based on underlying health conditions), transmission vulnerability (for example social mixing and hygiene infrastructure), health system vulnerability (for example hospitals beds and health workers etc) and vulnerability to control measures [72].

## Limitations

There were data related limitations when computing the vulnerability indices. Several variables such as access to mobile phones, access to insurance cover, and mobility between counties could not be accessed during the analysis. Further, due to the lack of granular data despite using SAE techniques, obesity, diabetes and hypertension could only be resolved at the county level. Therefore, we assumed the estimates within sub-county were equal for these three variables. Some of the datasets are not updated to 2020 and their trends are likely to have changed between 2014 and 2020. Further the analysis was conducted at the sub county level, it’s likely that that some variation and heterogeneity was masked in the relatively bigger polygons.

An equal weighting scheme was implemented for all the determinants. While variables can be weighted through epidemiological (amount of variation a variable explains in the outcome), subjective (from experts) or statistical approaches, there is neither a gold standard approach nor a predefined way of implementing the weights [51,53,73]. During the creation of an infectious disease vulnerability index [56], different weighting schemes were explored, and the results indicated the most vulnerable countries did not change substantially. However, the determinants employed in the current analysis are likely to have varying contribution, severity and impact to COVID-19 disease. The weighting scheme can be revised as more individual COVID-19 data with attributes become available in Kenya and other countries across Africa. This will allow a better definition of how the risks associated with different variables vary in across different population and settings [18].

The preliminary overlay between indices and the cases may have been limited. Cases were allocated in counties where they were recorded, however, these might not have been the residential counties for the last several years. Some might have been living outside Kenya and other counties. On the hand, the data layers used refer to the specific counties for the period between 2014 and 2020.

## Conclusions

Fighting the COVID-19 pandemic calls for precision public health reflecting our improved understanding of who is most vulnerable and what makes them more vulnerable, where the disease is spreading or likely to spread fastest, and where current interventions may not work as intended. COVID-19 is spreading at different rates across Kenya, most probably working its way through all 47 counties. The indices estimated presents tools that can be used by the Kenyan government and stakeholders to better plan especially in areas where COVID-19 has not been confirmed by prioritizing sub-counties that are moderately to highly vulnerable. The heterogenous nature of the vulnerability indices highlight the need to address social determinants of health disparities, strengthening of the health system and related programmes.

## Data Availability

All the datasets used in the current analysis are publicly available and can be accessed through references listed in Table 1 and in the supplementary files

## Declarations

### Funding

PMM is funded under the IDeAL’s Project, DELTAS Africa Initiative [DEL-15-003]. The DELTAS Africa Initiative is an independent funding scheme of the African Academy of Sciences (AAS)’s Alliance for Accelerating Excellence in Science in Africa (AESA) and supported by the New Partnership for Africa’s Development Planning and Coordinating Agency (NEPAD Agency) with funding from the Wellcome Trust [number 107769/Z/10/Z] and the UK government. PMM is also supported by funds provided under Professor RW Snow’s Wellcome Trust Principal Fellowship (#’s 103602 & 212176). EAO is supported as Wellcome Trust Intermediate Fellow (number 201866) that provided support for NJK; NJK, PMM and EAO, acknowledge the support of the Wellcome Trust to the Kenya Major Overseas Programme (# 203077). The views expressed in this publication are those of the authors and not necessarily those of AAS, NEPAD Agency, Wellcome Trust or the UK government. The funder of the study had no role in study design, data collection, data analysis, data interpretation, or writing of the report

### Authors’ contributions

All authors contributed to the development of the project, analysis, interpretation of the study findings and writing of the manuscript. All authors read and approved the final manuscript.

### Availability of data and materials

All the datasets used in the current analysis are publicly available and can be accessed through references listed in Table 1

### Ethics approval and consent to participate

This study used secondary data only, all publicly available and can be accessed from different data repositories via references listed in Table 1. No individual patient level data used in this publication.

### Consent for publication

Not applicable. The manuscript does not contain any individual person’s data.

### Competing interests

The authors declare that they have no competing interests

## Acknowledgements

The authors thank Professor Bob Snow for comments on data assembly and earlier versions of this manuscript

## Notes

### Competing Interest Statement

The authors have declared no competing interest.

### Funding Statement

PMM is funded under the IDeAL Project, DELTAS Africa Initiative [DEL-15-003]. The DELTAS Africa Initiative is an independent funding scheme of the African Academy of Sciences (AAS) Alliance for Accelerating Excellence in Science in Africa (AESA) and supported by the New Partnership for Africa's Development Planning and Coordinating Agency (NEPAD Agency) with funding from the Wellcome Trust [number 107769/Z/10/Z] and the UK government. PMM is also supported by funds provided under Professor RW Snow Wellcome Trust Principal Fellowship (number 103602 and 212176). EAO is supported as Wellcome Trust Intermediate Fellow (number 201866) that provided support for NJK; NJK, PMM and EAO, acknowledge the support of the Wellcome Trust to the Kenya Major Overseas Programme (# 203077). The views expressed in this publication are those of the authors and not necessarily those of AAS, NEPAD Agency, Wellcome Trust or the UK government. The funder of the study had no role in study design, data collection, data analysis, data interpretation, or writing of the report

